# Association Between Nonalcoholic Fatty Pancreatic Disease and Triglyceride/Glucose Index

**DOI:** 10.1101/2022.12.24.22283929

**Authors:** Luis Jesuino de Oliveira Andrade, Luis Matos de Oliveira, Alcina Maria Vinhaes Bittencourt, Gustavo Magno Baptista, Gabriela Correia Matos de Oliveira

## Abstract

Nonalcoholic fatty pancreatic disease (NAFPD) is an increase of fat in the pancreas, and has an important association with insulin resistance (IR) and type 2 diabetes mellitus. Research has confirmed that the triglyceridemia/glycemia (TyG) index determines IR as much as does the hyperinsulinemic-euglycemic clamp assessment as the homeostasis model testing of IR (HOMA-IR).

**Objective:** To evaluate the association between degree of NAFPD and TyG index.

**Methods:** In 72 patients undergoing ultrasound of abdomen with a diagnosis of NAFPD, insulin, glucose, and triglycerides levels were evaluated. The HOMA-IR index was used as a reference of IR. The degrees of NAFPD and the TyG index were presented through the receiver operating characteristics (ROC) curves in order to evaluate the association between different degrees of NAFPD and the TyG index and its correlation with HOMA-IR.

**Results:** There was a statistically significant correlation between the degree of NAFPD and the TyG index. The AUROC curve for the TyG index for predicting the degree of NADPD was 0.855 (0.840–0.865). The intensity-adjusted probabilities of the degree of NAFPD were more strongly associated with TyG values when compared with HOMA-IR.

**Conclusion:** In this study the TyG index correlated positively with the degree of NAFPD, performing better than HOMA-IR.

## INTRODUCTION

Pancreatic fat accumulation has been evaluated in research centers, and multiple terminologies and various concepts has been employed for this disease, particularly pancreatic fatty infiltration, pancreatic steatosis, lipomatous pseudohypertrophy, fatty pancreas, pancreatic lipomatosis, fatty replacement, nonalcoholic fatty pancreatic disease (NAFPD) and nonalcoholic fatty steatopancreatitis^1^. Currently, NAFPD has been the term used to describe the accumulation of fat in the pancreas^2^.

The pathophysiology of NAFPD begins with obesity. Thus, caloric excess will result in both hyperplasia and hypertrophy of adipocytes, with extravasation of triglycerides (Tg) into the peripancreatic tissue, leading to modifications in the ectopic fat microenvironment. The response to homeostatic changes results in the release of interleukins, tumor necrosis factor and macrophages, leading to receptor downregulation and inhibition in adipocyte differentiation^3^.

The TyG index has been used with a reliable marker for the diagnosis of insulin resistance (IR) as much as the hyperinsulinemic-euglycemic clamp test^4^, and when equated with the homeostasis model testing of IR (HOMA-IR)^5,6^. In 2010 Guerrero-Romero F, et al. published the manuscript “The product of triglycerides and glucose, a simple measure of insulin sensitivity. Comparison with hyperhyperinsulinemic euglycemic clamp”. In this study the TyG index was confirmed as a marker of IR through parity with the hyperhyperinsulinemic clamp and similarly to HOMA-IR^7^. However, the TyG index was proposed in 2008 by Simental-Mendia, et al.^8^, when they evaluated a population of healthy individuals in a cross-sectional study.

TyG index has been applied to healthy individuals as a marker of IR, as well as a marker of atherosclerosis and hepatic fatty infiltration^9,10^. However, there are no reports in the literature of the use of this index as a predictor of NAFPD. There are no relevant biochemical tests to assess pancreatic fatty infiltration. Thus, the aim of this study was to evaluate the association between the degree of NAFPD assessed by ultrasonography and the TyG index.

## METHODS

### Subjects and study design

The cross-sectional study included 72 individuals with NALPD diagnosed and graded using ultrasonography. The variables analyzed were age, gender, height, body mass index (BMI), neck circumference, abdominal waist, laboratory data (triglycerides, glucose, TyG index, insulin, HOMA-IR), and whole abdomen ultrasound.

The project was approved by the Conep (registry number: 2.464.513), carried out according to the provisions of the Declaration of Helsinki, and participants signed an informed consent.

### Biochemical Tests

Laboratory evaluation included determination of plasma insulin, triglycerides, and fasting glucose.

The fasting plasma insulin was determined by direct chemiluminescent technology (Access Immunoassay System), Triglyceride determinations were performed by the colorimetric enzymatic method, through a SELECTRA II Merck^®^ automated system, with reference values lower than 150 mg/dL. Fasting glycemia was measured by the colorimetric enzymatic method (GOD-PAP) using as reference values the Brazilian Guidelines of the Brazilian Society of Diabetes 2022^11^.

### Estimation of biochemical parameters

The HOMA-IR was calculated using the fasting glucose level (mmol/L) multiplied by fasting insulin level (lU/mL) and then divided by 22.5^12^.

The TyG index has its calculation based on the lipid profile, using the serum Tg value, and on fasting glycemia, and its formula is TyG = Ln [Tg (mg/dL) x Glycemia (mg/dL)/2], in which Ln is the neperian logarithm^8^.

### Pancreatic ultrasound examination

Ultrasonographic evaluation of the pancreas was performed the same expert radiologist, who were blinded to biochemical and clinical results, and evaluated the findings of pancreatic ultrasound examination using the FIGLABS FT412^®^ equipment, with a 3.5 MHz frequency convex transducer, through direct contact. Pancreatic steatosis was classified into four grades, according to the echogenicity of the pancreas in relation to the renal echogenicity and the echogenicity of the retroperitoneal fat^13^.

### Statistical analysis

Data were analyzed using the public domain statistical program R 3.1.1. To consider a variable having normal distribution the following parameters were used: mean, median, standard deviation, symmetry and flattening of the curve, histogram, Q-Q plots and Kolmogorv-Smirnov test for normality. We performed uni and multivariate logistic regression analyses to define the impact of various TyG levels on NAFPD risk. The potential of the TyG index in predicting NAFPD was examined by receiver operating characteristic (ROC) curve analysis and area under the curve (AUC) values.

Values of P < 0.05 (5 %) were adopted as significance level.

## RESULTS

### Demographic characteristics

Data from 72 patients were included, 51.40% male and 48.60% female. The subjects had a mean age of 45.31 ± 13.72 years, weight 88.52 ± 18.27 kilos, BMI 32.25 ± 5.73 Kg/m^2^, and waist circumference (WC) of 99.68 ± 14.52cm. The data are presented in Table 1.

**Table 1.** Demographic characteristics of the study population

| Factor | Minimum | Median | Average | Maximum | Standard Deviation |
| --- | --- | --- | --- | --- | --- |
| Age (years) | 20 | 43.0 | 45.31 | 65 | 13.72 |
| Weight (Kg) | 45.00 | 86.00 | 88.52 | 125.00 | 18.27 |
| Height (m) | 1.47 | 1.64 | 1.66 | 1.89 | 10.21 |
| BMI (Kg/m<sup>2</sup>) | 19.90 | 30.80 | 32.25 | 47.50 | 5.73 |
| WC (cm) | 52.00 | 99.50 | 99.68 | 140.00 | 14.52 |

The prevalence ratio between normal BMI, overweight (OW), grade I obesity (OG1), grade II obesity (OG2), grade III obesity (OG3) and NAFPD was 14.30%, 50.00%, 75.00%, 57.10 and 66.70% respectively. The estimated risk (OR) in the sample of the subject with normal BMI, OW, OG1, OG2 and OG3 develop NAFPD was 0.098 (0.011 - 0.860), 0.625 (0.221 - 1.767), 3.286 (1.161 - 9.296), 0.941 (0.289 - 3.064) and 1.450 (0.125 to 16.786) respectively (Table 2).

**Table 2.** Association between Weight and NAFLD

| Association | Prevalence Ratio (%) | Estimated Risk (OR) |
| --- | --- | --- |
| **Normal weight / NAFLD** | 14.30 | 0.098 (0.011-0.860) |
| **Overweight / NAFLD** | 50.00 | 0.625 (0.221-1.767) |
| **Grade I Obesity / NAFLD** | 75.00 | 3.286 (1.161-9.296) |
| **Grade II Obesity / NAFLD** | 57.10 | 0.941 (0.289-3.064) |
| **Grade III Obesity / NAFLD** | 66.70 | 1.450 (0.125-16.786) |

### TyG index and NAFPD

The Fasting glucose (mg/dL), Triglycerides (mg/dL), TyG index, and HOMA-IR associated with NAFPD were 120.71 ± 66.66, 227.5 ± 175, 4.95 ± 0.37, and 2.88 ± 2.10 respectively (Table 3).

**Table 3.** Association between Fasting glucose, Triglycerides, TyG and NAFLD

|  | NAFLD | P value |
| --- | --- | --- |
| Fasting glucose, mg/dL | $120.71 \pm 66.66$ | 0.02 |
| Triglycerides, mg/dL | $227.5 \pm 75.01$ | 0.01 |
| TyG index | $4.95 \pm 0.37$ | 0.001 |
| HOMA-IR | $3.40 \pm 2.06$ | 0.08 |

A higher TyG index, TyG-BMI, TyG-WC were correlated with a higher degree of NAFPD. Estimated Risk for NAFPD increased in all indices, particularly in TyG-BMI. The area under the receiver operator characteristic curve (AUROC) was 0.855 (0.840–0.865) for TyG-NAFPD (Figure 1).

**Figure 1.**
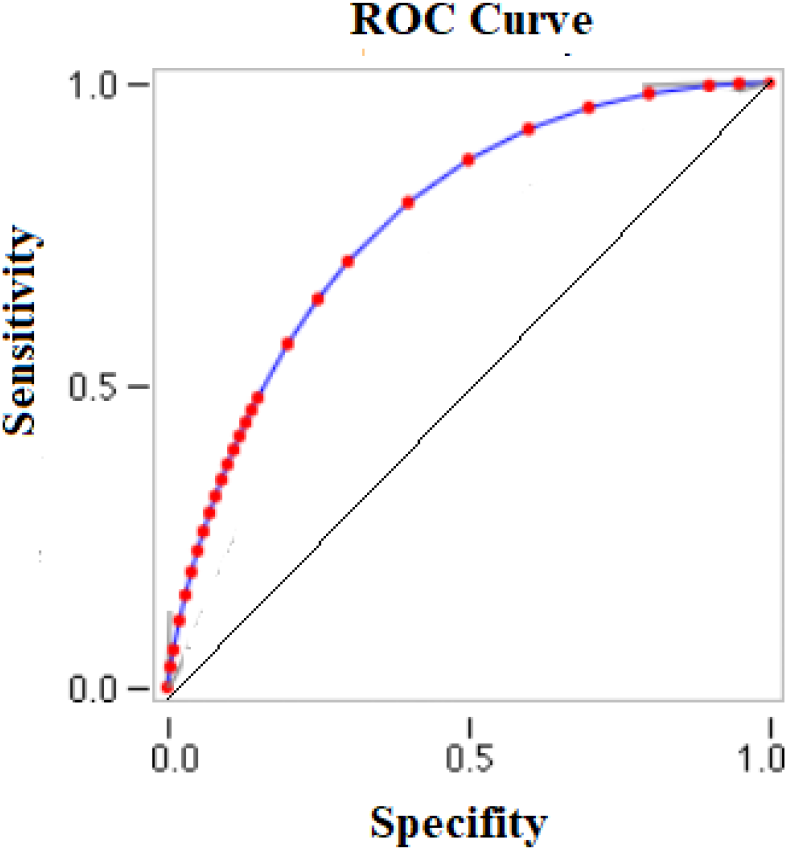
Curve for the prediction of NAFPD by the TyG index.

The adjusted probabilities of NAFPD grade intensity were more strongly associated with TyG values when compared with HOMA-IR, as described further below.

### HOMA-IR and NAFPD

The probability of NAFPD was the highest when HOMA-IR was above 3.1 and triglyceride above 206 mg/L. This combined model resulted in a higher AUROC curve (0.80), than HOMA-IR individually.

## DISCUSSION

We evaluated the association between the TyG index and NAFPD which showed a high association. We did not find any studies evaluating the association between TyG and NAFPD in the literature between them.

Studies suggest that serum triglyceride assessment should be incorporated as a clinical and laboratory indicator of NAFPD^14^. The TyG index has been associated with metabolic syndrome, hepatic steatosis, and cardiovascular disease^15^. However, it is unclear whether the TyG index is associated with NAFPD, however as this index is associated with IR consequently it is deduced that the TyG index is related to NAFDP.

In addition to the TyG index and HOMA-IR for IR assessment, the Matsuda index and the Raynoud index have been described^16,17^. However, the gold standard for IR assessment is the hyperinsulinemic-euglycemic clamp test^4^. We evaluated the association between the TyG index and NAFPD, also comparing it with HOMA-IR.

Pancreatic fatty infiltration is an expression to describe adipocyte infiltration inside of pancreatic parenchyma, instead replacement by fatty tissue, and is linked to obesity and metabolic syndrome^18^. Thus, NAFPD is a significant complication of obesity, may be an important factor for pancreatic complications resulting from obesity. Our study demonstrated a significant association between the degrees of obesity with the degree of NAFPD, with an estimated risk three times higher than in individuals with normal BMI.

Unesterified fatty acid stimulates insulin secretion leading to hyperinsulinemia and consequently IR^19^. Recent study and experimental data showed that pancreatic fat cells lead to degeneration of pancreatic beta cell function only in a typical environment characterizing IR^20^. Thus, NAFPD presents an important correlation with the beta cell dysfunction, IR and reduced insulin secretion. Our results showed correlation between NAFPD and IR assessed by HOMA-IR; however TyG index showed a better correlation with NAFPD when compared with HOMA-IR.

NAFPD is an important challenge in medicine and should be taken into consideration as an important risk factor for IR and its consequences, although it is questionable whether NAFPD is really the cause of IR or part of it. Thus, it is important to correlate NAFPD with markers of IR, especially the TyG index, which in our study showed a significant correlation.

## CONCLUSION

In this study the TyG index correlated positively with the degree of NAFPD, performing better than HOMA-IR. However, new studies to evaluate this correlation, as well as multicenter trials should be conducted in this area, using the TyG index because it is an easy-to-perform, less expensive, and clinically validated method.

## Data Availability

All data produced in the present work are contained in the manuscript

